# Comparison of treatment planning techniques at the linear accelerator for left-sided breast carcinoma considering the treatment volume and organs at risk

**DOI:** 10.1101/2025.10.28.25338144

**Authors:** Marvin Christopher Stegmann, Kay C. Willborn, Jürgen Trzewik, Sabina Stroh, Maximilian Even, Jil Dailin Thiemann, Frederik Morgenroth, Björn Poppe

## Abstract

Breast cancer is the most common type of cancer in women. Nowadays, postoperative radiation therapy is one of the standard treatment procedures. However, the radiation used may not only lead to a positive therapeutic outcome, but also has a negative impact on the surrounding organs at risk. Organs, such as the heart, can also have damaged and serious late effects can occur. In recent decades, there has been a significant development in the field of equipment technology and radiation planning. Today various radiation planning techniques are available, such as 3DCRT (three-dimensional conformal radiation therapy), VMAT (Volumetric Intensity Modulated Arc Therapy) or IMRT (Intensity Modulated Radiation Therapy). The question arises as to which of the radiation techniques is most suitable for breast cancer radiation. In this study, the radiotherapeutic treatment of left-sided breast carcinoma using linear accelerators has been investigated. The radiation techniques 3DCRT, HYBT (hybrid technique [3DCRT+VMAT]), VMAT and IMRT were compared. In addition, the breast volumes/shapes were divided into breast groups in order to show a possible dependence on the target volume. CT images of 20 different patients were used; the patients were divided into four breast groups. All irradiation techniques were applied to the same CT. A total of 80 radiation plans were created and compared. In the evaluation, the mean dose or max dose values, the MU count of the irradiation plan, the breast group, the homogeneity index and the conformity index were considered, depending on the organs at risk. 3DCRT has historically been the most widely used technique. It has been found that 3DCRT normally provides a better sparing of the dose to OAR, when compared to newer techniques. Regardless of the radiation techniques, for technical reasons, complete protection of the organs at risk, such as the heart, cannot always be achieved even when the planning system is fully utilized. This is due to anatomical reasons, as the chest practically wraps around the thorax in the supine position.

## 1. Introduction

The most common carcinoma in women in Germany is breast carcinoma at 29.5%. Taking current statistics into account, one in eight women will develop breast cancer. Three out of ten patients are under the age of 55, which makes it even more important to minimize the damage to healthy, normal tissue. [1]

Today, postoperative radiation therapy of the breast is the standard treatment for breast cancer, along with surgery, chemotherapy and hormone therapy, and yet the use of ionizing radiation can not only lead to a positive therapeutic outcome, but also carries certain risks. The radiation can damage surrounding tissue and organs, which can lead to several unwanted side effects. [2] [3]

Organs at risk associated with breast radiation therapy may include the heart and coronary arteries, the RIVA (Ramus interventricularis anterior) or the lungs. Radiation damage to the coronary vessels has been shown to lead to cardiovascular damage and a resulting significantly increased likelihood of suffering a heart attack up to 15 years later. The lung is also an OAR (Organs at Risk) near the Planning target volume (PTV). Radiation damage to the lungs leads to lung fibrosis and the risk of developing lung cancer increases significantly in the years following radiation[4], [5], [6], [7], [8] [9]. In addition, the healthy contralateral breast should also not be neglected when considering the organs at risk. The risk of a second carcinoma increases to 2.5 times the risk even at low organ doses of > 1.0 Gy [10]. In this study the following OARs are considered due to their proximity to the high dose area of PTV: heart, left lung, RIVA and the contralateral breast. Other anatomically distant OARs, such as the spinal cord, will also be addressed later.

In this study, the radiotherapeutic treatment of left-sided breast carcinoma using linear accelerators was investigated.

The comparison of different irradiation techniques for breast irradiation has been discussed for some time by various research groups, but no clear statement can be made due to different results. For example, Rastogie et al. showed that IMRT is advantageous compared to 3DCRT regarding high-dose areas of the ipsilateral lung and heart. With regard to low-dose areas, 3DCRT again seems to be superior to IMRT. [11] Another study was conducted by Das Majumdar et al. comparing 3DCRT, IMRT and VMAT in post-mastectomy patients. This study also concluded that both IMRT and VMAT showed better results in terms of dose distribution compared to 3DCRT for high dose irradiation. In case of low-dose irradiation, 3DCRT again proved to be advantageous. [12] Garg et al. (2022) compared the IMRT and 3DCRT in breast cancer irradiation in terms of dose to the heart and concluded that 3DCRT gives a significantly lower dose to the RIVA and the heart. However, the maximum dose to the RIVA and the heart showed no significant differences between the two radiation techniques. [13] Hall et al. (2003) showed in their study that 3DCRT provides better results compared to IMRT for breast cancer irradiation. IMRT would approximately double the incidence of second malignancies due to damage to the OAR. [14]

The S3 guideline for breast cancer [2] currently recommends that IMRT or VMAT should not be used for adjuvant radiotherapy of breast cancer in general, but only for larger breasts and/or abnormal thoracic curvature, such as a funnel chest, due to the anatomical conditions and the resulting increased distance from the irradiated area to organs at risk. [15], [16], [17] IMRT can achieve increased dose homogeneity, but usual not beneficial organ conformity.[16], [18] Patient positioning techniques such as deep inspiration can additionally contribute to a lower OAR exposure of the heart. [18]

To take up the results of previous studies, as well as the recommendation of the S3 guideline for breast carcinomas, and to investigate to what extent these data can be confirmed, this study also examines the different radiation techniques regarding the avoidance of normal tissue. The aim of the study was to apply the different irradiation techniques to defined breast groups and to investigate whether the different breast sizes, as mentioned in the S3 guideline for breast carcinomas, have an influence on the results. Due to the high risk of the heart as an OAR, the heart exposure of the patients was particularly considered in relation to the different breast volumes.

## 2. Materials and Methods

This is an experimental clinical research based on existing patient data. For this purpose, computed tomographic thorax images of 20 different patients were selected. The CTs of the patients were divided into four groups and for each patient a radiation plan without validation of regional lymph nodes was calculated using all four radiation techniques. Thus, a total of 80 radiation plans were generated, evaluated and compared. The patients were all positioned on the same patient positioning board (Kombiboard Bj. 2013, Fa. Unger, Germany), with similar board settings, i.e. with the arms above the head and without inclination of the lying surface. The bar, which prevents the longitudinal displacement of the patients, was individually adjusted to the patient’s size. All participating patients were able to maintain this position during radiation. The CT images were all acquired in “head-first-supine” position with a slice thickness of 2 mm. The evaluation focused primarily on the PTV. The PTV (Planning Target Volume) was plotted by an authorized radiation therapist, according to the current specifications of the Contouring Atlas of the RTOG (Radiation Therapy Oncology Group Journal of Applied Clinical Medical Physics, Volume 15, Number 2, 2014). The radiation planning program “Eclipse” from Varian, version 15.6, was used for the calculation. The planning was based on irradiation data from a Varian Truebeam linear accelerator (year 2013). Photon energies of 6 MV and 15 MV were available for the irradiation planning. The data required for evaluation were taken directly from the planning system.

**Table 1:** breast group overview.

| Label | Abbreviation | Volume |
| --- | --- | --- |
| **Standard breast** | Std | 700 – 1000 cm<sup>3</sup> |
| **Small breast** | K | < 700 cm<sup>3</sup> |
| **Large breast** | G | > 1000 cm<sup>3</sup> |
| **Thoracic wall** | Thxw | State after mastectomy |

Only patients with left-sided breast carcinoma were included in the study. In addition, the patients were independently classified into different breast groups (small, large and standard breasts) based on average breast volume of the of the available participants. Thoracic walls resulting from a mastectomy were added regardless of volume, as in these cases the thin target volume was of particular interest regarding radiation planning. In addition to the wall thickness, the volume is also very strongly dependent on the associated longitudinal target volume length. This varies due to the poorly definable irradiation volume. The following breast groups were defined: Standard breast (Std, 700 - 1000 cm^3^), small breast (K, < 700 cm^3^), large breast (G, > 1000 cm^3^), thoracic wall (Thxw, S/P. masectomy). The size of the patient was not taken into account, only the pure volume of the breast.

The irradiation techniques used were 3DCRT, IMRT, VMAT and a hybrid technique of 3DCRT and VMAT (HYBT) [19]. In 3DCRT, two large opposing main fields were initially created, which completely cover the target volume and through which a large part of the dose is applied. To properly cover the movement of the breast as it swells during therapy, the fields were extended 30 mm outward from the skin. The irradiation fields were applied tangentially to the thorax and an MLC was used for individual modulation to the target volume. Subsequently, additional dose saturation fields (subfields) were added in order to achieve a homogeneous dose distribution in the target volume.

The IMRT planning was performed using the sliding window technique. The irradiation fields were not placed directly opposite each other. The offset to the opposite field was at least five degrees. Then, the constraints for the OAR were defined and adjusted after the first optimization step. To ensure a homogeneous dose distribution in the target volume, the tool “Skin Flash” from Varian was used. Here, the leaves were opened 30 mm anteriorly from the breast surface to prevent a small opening between the leaves and to consider the additional movement of the target volume. In the next step, the fluences were observed and hotspots were homogenized using “Smooth Transmission Factor” and missing dose was added using the “Increase Transmission Factor” tool.

By selecting appropriate start and stop angles, the VMAT irradiation plans were designed to minimize radiation through the OARs indicated above and to overlap them with the contour of the PTV near the thoracic wall. The collimator angle was then adjusted to follow the contour of the PTV towards the lung. After the first optimization with the corresponding constraints, the rarely used angles were then removed from the available irradiation angles in the control points (angle-dependent specification of the gantry speed and number of MU) in a new optimization, so that these were not used in the optimization and mostly fan-shaped arcs were created. This meant that small, hardly used irradiation fields could be avoided.

For the hybrid technique (HYBT), the 3DCRT and VMAT techniques were combined (see Figure 1). First, two main arcs were created similar to the 3DCRT planning, normalized to 80 % of the prescribed dose and the missing dose was saturated using VMAT. For this purpose, fan-shaped arcs were directly created without having previously performed an optimization. The previously created 3DCRT plan was specified as the “base dose plan” in the optimization. All inverse planned radiation techniques were optimized using the following OARs: heart, lung left, lung bilateral, RIVA (Ramus interventricularis anterior).

**Figure 1:**
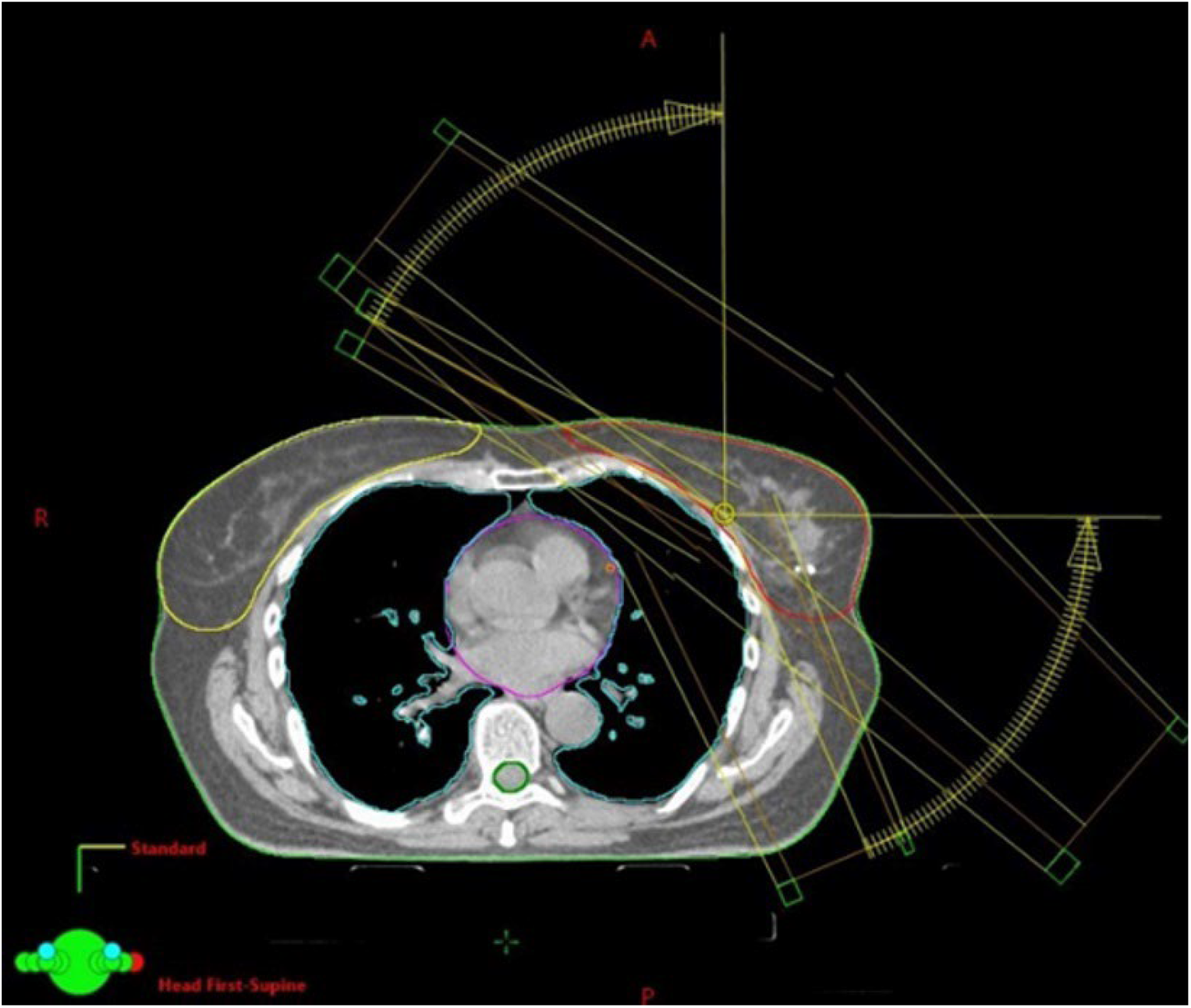
Transverse cross-section of a HYBT with 3DCRT irradiation field and fan-shaped arc segments

For the evaluation, the data calculated by the planning system were exported and further processed using a spreadsheet program. The conformity index (CI) and the homogeneity index (HI) were calculated. To ensure a critical view and to consider the total radiation dose in the body, the CI considered the coverage of the PTV in consideration of the isodose in the whole body and not only in the target volume. The HI was calculated for the individual breast groups and for a mean value formed from the breast groups depending on the irradiation technique.

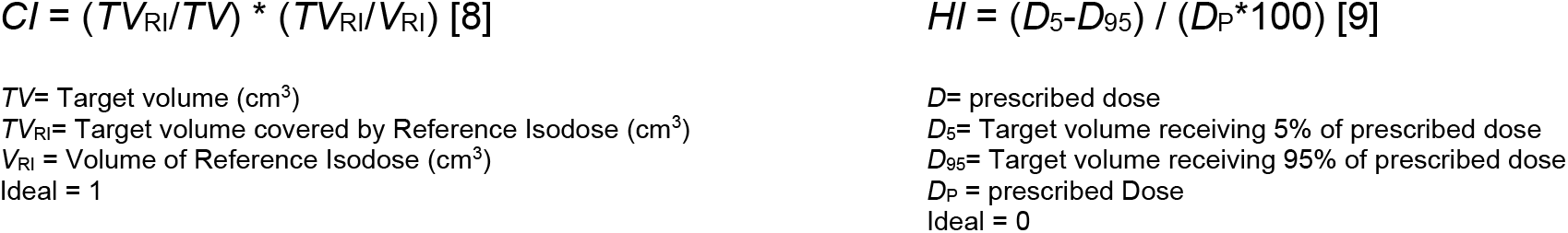

Furthermore, the maximum monitor units (MU) of the irradiation plan, the breast volume and the mean and maximum values of the respective OAR were read from the system. Mean values (M) and standard deviations were calculated. The following OARs were considered: heart, left lung, RIVA (Ramus interventricularis anterior) and the contralateral mamma.

## 3. Results

In order to be able to make a statement about the dose exposure of the **organs at risk** typical in breast cancer irradiation, the dose-mean values of the OARs located in the immediate vicinity of the PTV and the dose-max values of the OARs located further away from the PTV were examined.

The **dose-mean values** of the left lung (L), the RIVA, and the contralateral mamma were considered without taking into account the breast group. (see Figure 2). Due to the severity of possible side effects of radiation exposure to the heart, a plot with a breakdown of the breast groups was created for this purpose to show the influence of the different breast groups on the dose in this OAR (see Figure 3). On average, the heart shows the highest exposure with VMAT (3.5 Gy); followed by IMRT (2.7 Gy), HYBT (2.5 Gy) and 3DCRT (1.7 Gy). A similar burden depending on the irradiation technique can be observed between the different breast groups. 3DCRT achieved the best result in all breast groups.

**Figure 2:**
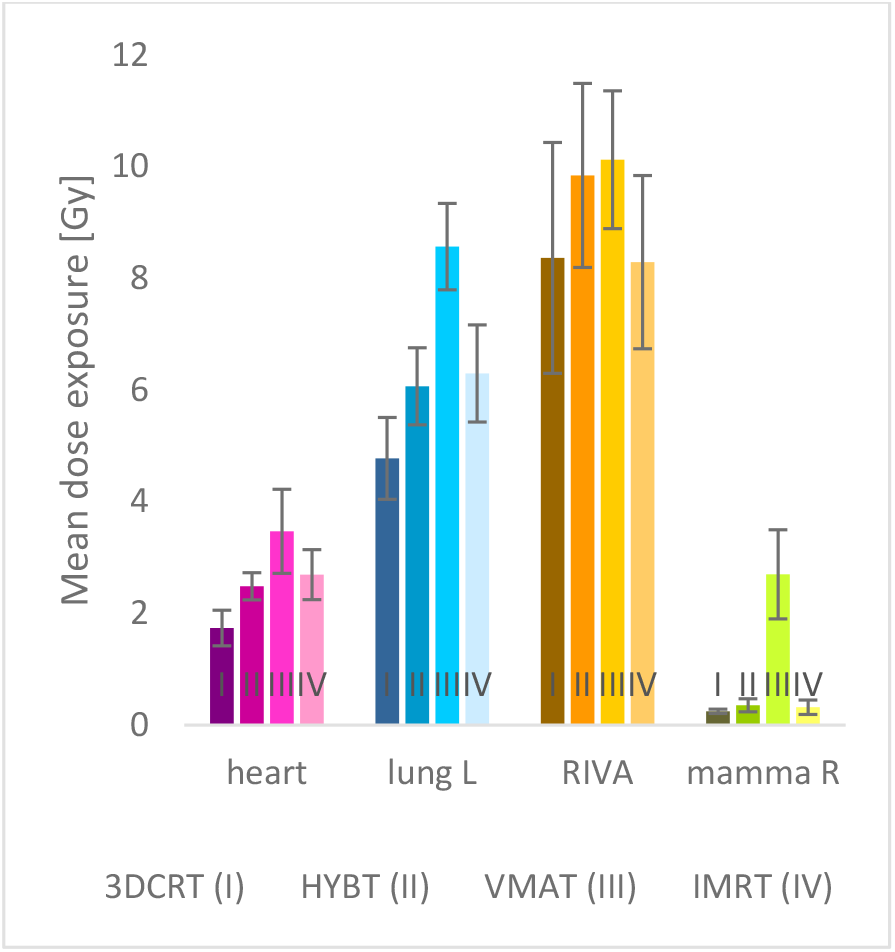
Mean dose exposure of the OAR

**Figure 3:**
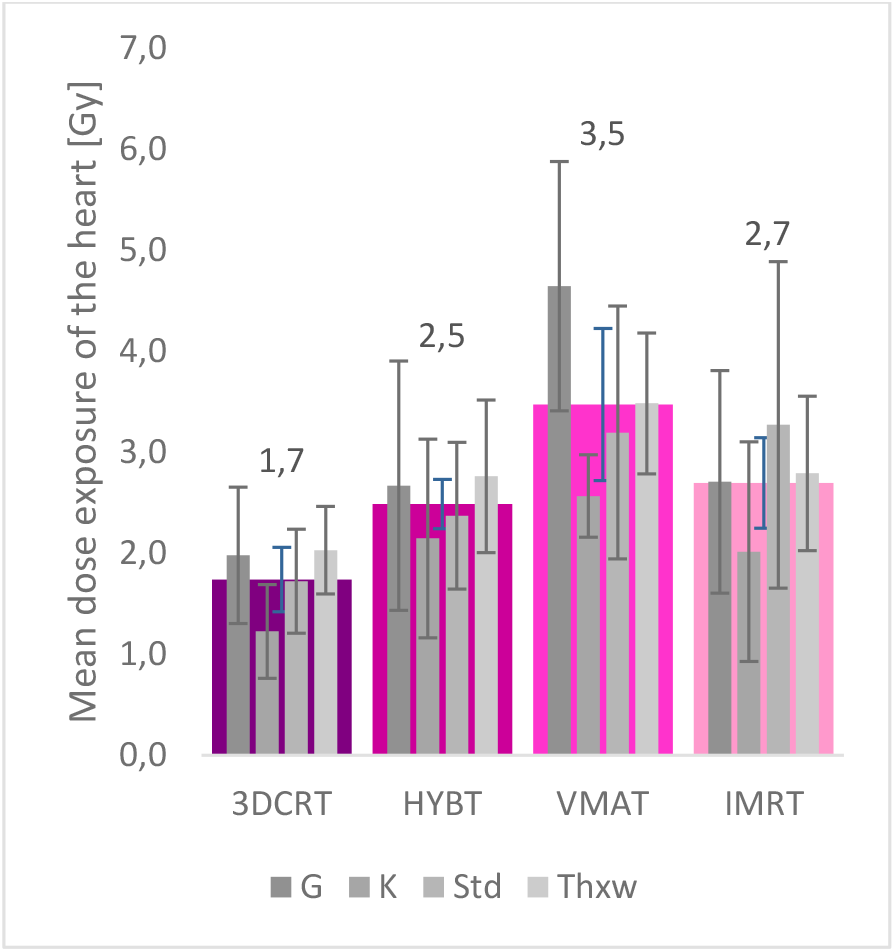
Mean dose exposure of the heart

In Figure 2, VMAT shows by far the highest exposure in the **contralateral mamma** (mamma R) compared to the other techniques. 3DCRT, HYBT and IMRT show a similar exposure of the contralateral mamma. When considering the RIVA, 3DCRT, IMRT, HYBT as well as VMAT perform similarly, with HYBT and VMAT showing the highest mean dose values for the **RIVA**. The ipsilateral **lung on the left** receives the lowest dose with 3DCRT, and the highest exposure is seen with VMAT. IMRT and HYBT show a similar value.

In the assessment of the OARs with greater distance (see Figure 4), the spinal cord, the right lung (lung R) and the contralateral mamma were considered regarding the **dose maximum values**. The individual breast groups were not considered separately. Compared to the other techniques, VMAT has the highest dose for all OARs considered. The exposure of the contralateral breast is similar for 3DCRT, HYBT and IMRT (approx. 7.6 Gy). A significantly higher exposure is observed for VMAT (approx. 16.5 Gy).

**Figure 4:**
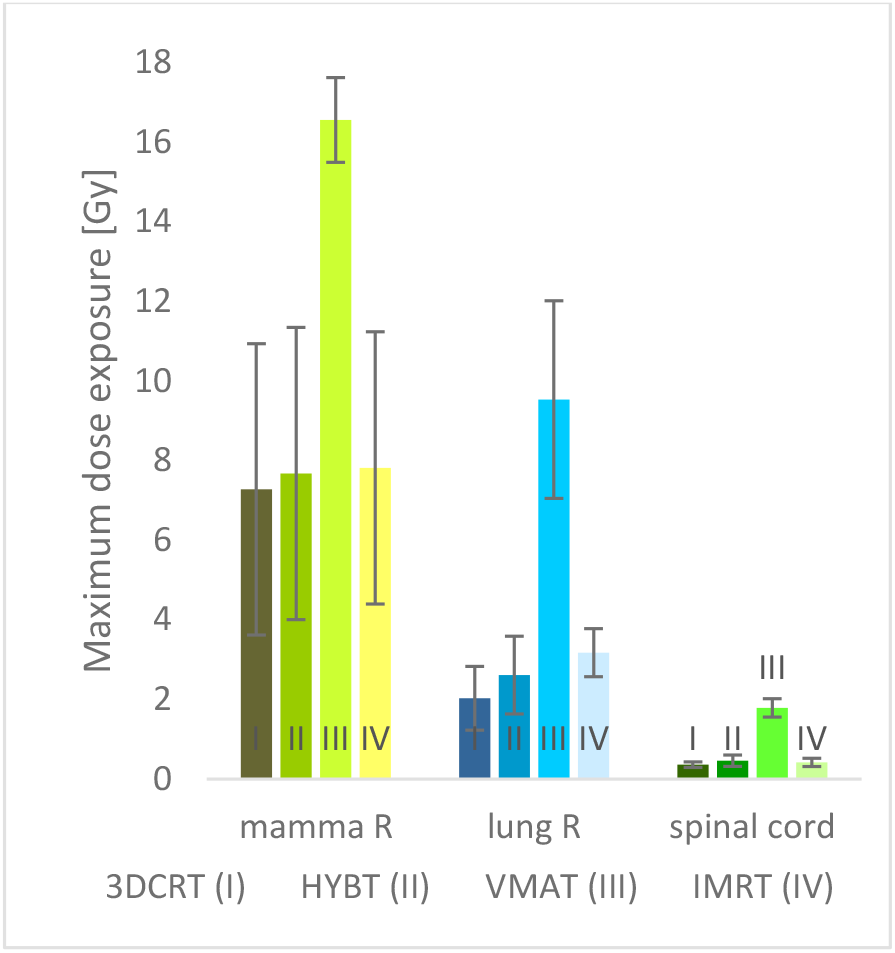
Maximum dose exposure of the OAR

The spinal cord is similarly exposed with 3DCRT, HYBT and IMRT (approx. 0.41 Gy). With VMAT, the exposure is more than four times higher at 1.8 Gy. The lungs receive a significantly higher dose of 9.5 Gy with VMAT compared to 2.0 Gy with 3DCRT, 2.6 Gy with HYBT and 3.2 Gy with IMRT.

### Monitor units (MU)

There are differences in the number of monitor units between the individual irradiation techniques (see Figure 5). IMRT has by far the highest number of MU (average approx. 904 MU), followed by VMAT with 368 MU. With 230 MU, 3DCRT requires the least MU. HYBT, with 255 MU, falls between the 3DCRT and VMAT techniques.

**Figure 5:**
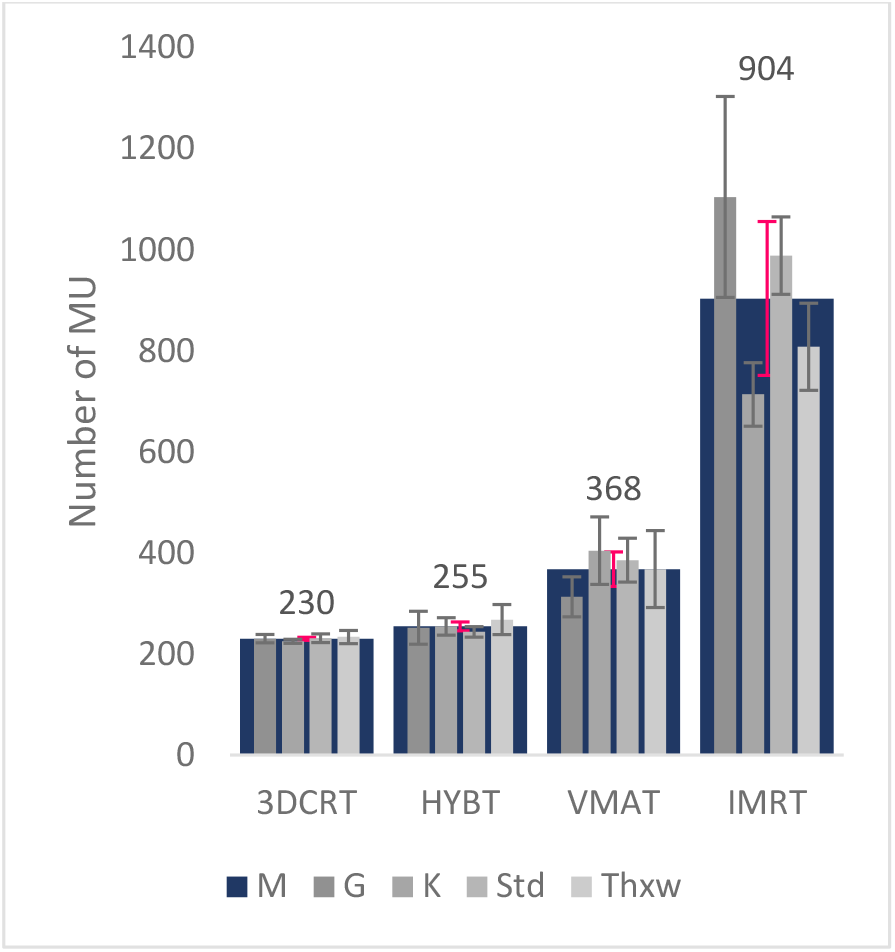
Comparison of the MU

The number of MU under consideration of the breast groups is very similar within the respective irradiation technique, larger differences can only be observed with IMRT.

### Homogeneity index

The homogeneity index is lowest for IMRT with 10.8 (see Figure 6), followed by VMAT with 12.4, HYBT with 13.1 and 3DCRT with 14.1. HYBT lies between 3DCRT and VMAT in the mean value. When looking at the individual breast groups, it is noticeable that breast group G has a more homogeneous distribution than the mean value of the respective irradiation technique. When looking at the individual breast groups, it is evident that the breast group Std has a high HI compared to the others for all techniques. The K and Std groups always have a higher HI than G and Thxw. The K group only has a lower value for VMAT.

**Figure 6:**
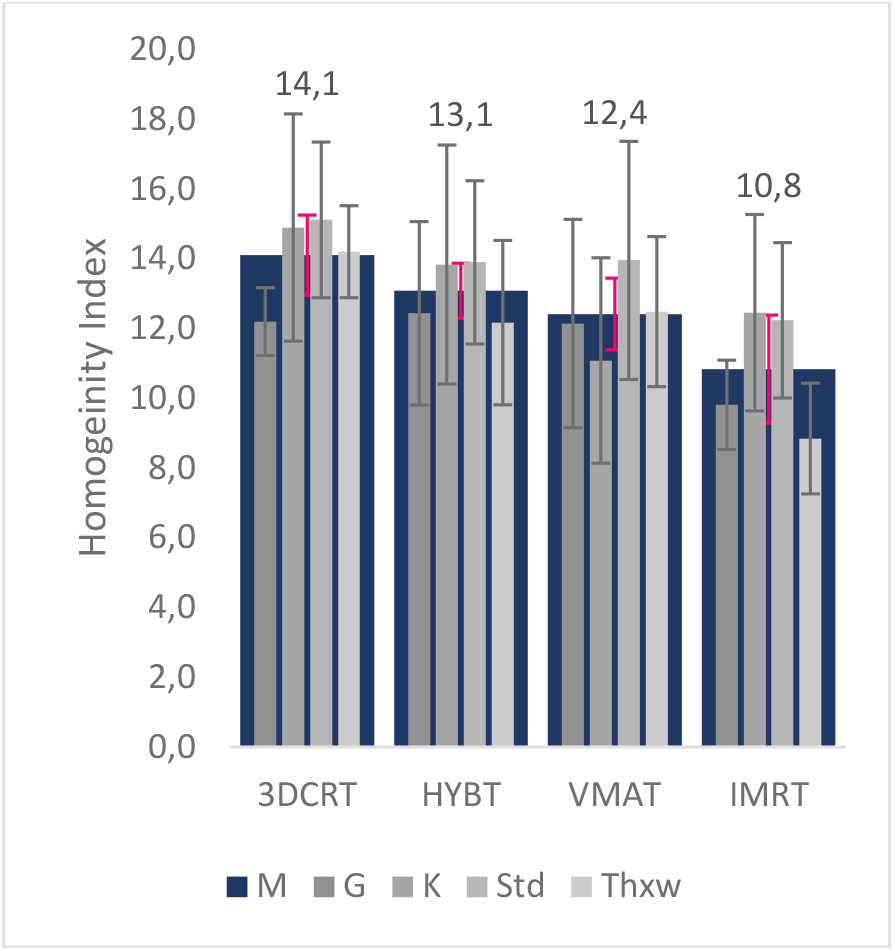
Comparison of the HI

### Conformity index (CI)

The coverage of the PTV was analyzed in consideration of the prescribed isodose in the whole body (see Figure 7). VMAT has the highest CI (0.82) on average for all breast groups considered, followed by IMRT (0.74), HYBT (0.73) and 3DCRT (0.69). In this case, HYBT lies between 3DCRT and VMAT. IMRT and HYBT have a similar mean value. When looking at the individual breast groups, group G has the highest CI for all techniques. Larger breast groups show a higher CI.

**Figure 7:**
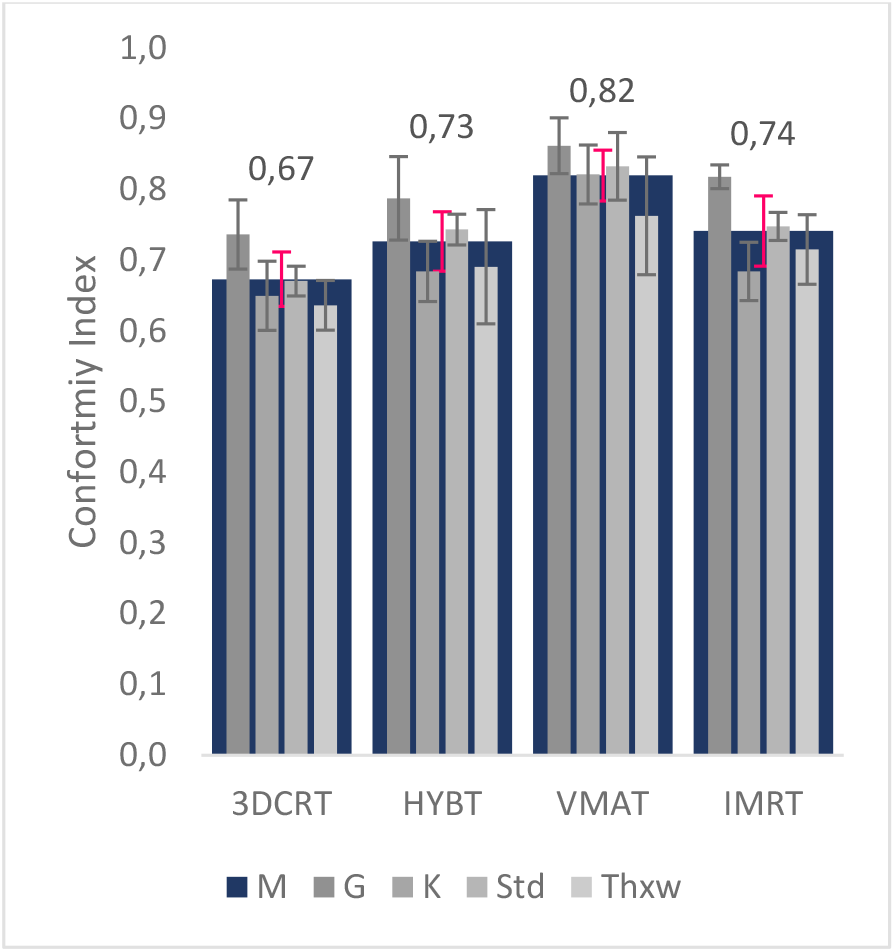
Comparison of the CI

## 4. Discussion

The **dose-mean values** of the risk organs located in the direct proximity of the left breast were considered. The dose-mean values of individual organs such as the heart are to be taken into account in addition to the concrete dose-volume data when assessing the risk organ exposures.

### Heart

The mean value is very important when assessing the dose to the heart. According to Piroth et al. this should be less than 2.5 Gy when considering the entire heart. Nevertheless, apical areas of the heart such as the RIVA and the left ventricle receive significantly higher doses than 2.5 Gy. The doses in this case are < 3 Gy for the left ventricle and < 10 Gy for the RIVA. For the assessment of the cardiac dose, the mean values of the entire heart are still referred to. In the 1970s to 1990s, significantly higher mean cardiac exposures were accepted, these were 13.3 Gy (1970) and 4.7 Gy (1990) [9]. According to our results, only HYBT and 3DCRT are equal to or below 2.5 Gy and proved to be more beneficial in terms of sparing the heart. These results can be compared with those of Garg et al. who also concluded that 3DCRT is the least stressful in terms of dose-mean values to the heart. IMRT and VMAT are 2.7 Gy and 3.5 Gy, respectively, but still within an acceptable range. It can be assumed that this comes from the different directions of irradiation. IMRT with its angled fields and VMAT with its direct beams to the center of the body have more dose modulation possibilities due to the different beam directions and can thus achieve a better dose distribution. This is also confirmed by the results of dose homogeneity; these two techniques provide the most homogeneous results. However, at the same time they lead to an increase in the mean dose on the hearts. The mean dose load to the hearts is lowest for small breasts, regardless of technique, due to the often compact shape and attached position to the thorax. In relation to this, larger breast volumes behave differently in the supine position; they are positioned more around the thorax and thus also more around the heart. There are major differences in the choice of irradiation technique. Despite the limited modulation options, 3DCRT achieved the best result in all breast groups. This can be explained by the typical target volume structure, as the target volume is tangential to the body and is not radiated directly in the center of the body in 3DCRT.

Thoracic walls are the result of a complete mastectomy and are usually very narrow irradiation volumes directly adjacent to the ribs. Delineation of the original breast tissue is often not possible after surgery and therefore these volumes are often defined with a wider safety margin. The target volumes often extend over large parts of the thoracic side and therefore automatically lie like a wreath around the heart in a near-thoracic cardiac position, making treatment planning much more difficult. In comparison to the irradiation techniques used, the use of electron radiation in Thxw would have been interesting. Because of the shorter range in matter, electron radiation is not recommended for all chest irradiations. [20], [21] Nevertheless, its use in the irradiation of the Thxw would be possible due to the lower PTV thickness. In this case, the appropriate dose coverage at different energies as well as the dose of OAR would need to be further investigated. [20], [21]

### Contralateral breast

The contralateral mamma should be considered as an OAR during treatment planning, both when creating the field position and during optimization. A low dose (>1 Gy) may be sufficient to produce a second carcinoma in the contralateral breast. Especially women younger than 40 years have a 2.5 times increased long-term risk with such a dose [10]. Therefore, the dose in this area should also be kept as low as possible [22].

VMAT clearly shows the highest dose-mean-load in the contralateral mamma. Compared to the other techniques, it is about 9 times higher on average. 3DCRT has the lowest dose with a mean value of 0.25 Gy. The large differences can be explained by the characteristics of the individual irradiation techniques. In 3DCRT, the irradiation fields were designed in such a way that the tangents cover the trailing edges of the breast target volume, but spare the OAR as much as possible. Among other things, it was ensured that the main fields did not extend too far into the contralateral mamma. For this purpose, the sternum ending in the contralateral mamma was used as a reference point. For IMRT, the irradiation fields were set up similarly to 3DCRT; again, extra consideration was given to the contralateral mamma, with the opposing fields extending further into the contralateral mamma than in 3DCRT. Beckham and Lohr et. al. describe the low-dose behavior of IMRT very similarly. More complicated target volumes can be better achieved while sparing critical organs, but not while avoiding an increase in the total dose to normal tissue [16], [17]. In VMAT, larger arc segments had to be selected, some of which were also aligned through the contralateral mamma, in order to provide the planning program with sufficient modulation options. The contralateral mamma was also considered as OAR in the optimization. In total, however, significantly more radiation is delivered through the contralateral mamma in VMAT, which explains the increased mean dose. If necessary, the OAR volume should have been considered more in the optimization in order to achieve a similar result. Unexpectedly, the HYBT did not place itself exactly between the 3DCRT and the VMAT, as it did with the other results, but also had a low dose. This can be explained by the weighting; the 3DCRT portion was dosed at 80% and the VMAT portion at 20%. Thus, the proportion of low exposure to 3DCRT is significantly higher and in total, the additional VMAT does not lead to a significant dose increase in the contralateral mamma.

When looking at the **RIVA**, no significant differences are observed. HYBT and VMAT have slightly higher dose values than 3DCRT and IMRT. On average for all irradiation techniques, the RIVA received a dose of 9.2 Gy.

Similar to what Rastogie et al. found, the **lung L** adjacent to the target volume showed the greatest sparing with 3DCRT. [11] HYBT and IMRT behaved similarly with loading. The increased load with VMAT was conspicuous. Yet again, as already considered for the contralateral mamma, the increased dose can be attributed to the characteristics of the irradiation techniques. Compared to 3DCRT, VMAT radiates significantly more into the body and thus naturally also increases the dose to the OAR located there.

For an even better estimation of the dose load in the body with the different irradiation techniques and, if necessary, to be able to draw conclusions about a possible radiation-induced second carcinoma, the **dose-max values** of more distant OAR (contralateral mamma, lung R, spinal cord) were taken into account. The dose-max values considered are very low. However, it was shown that even low dose values are relevant when considering organs at risk. [10]

According to the first results, the VMAT is expected to have the maximum dose exposure in all three OARs considered. The maximum dose to the **contralateral breast** is twice as high compared to the mean of the other three techniques. In the lung R, 3DCRT is the best, compared to the mean of the other three techniques, the exposure from VMAT is 3.7 times higher; in the spinal cord it was 4.3 times higher.

### Monitor units

The number of monitor units is significantly higher for IMRT than for the other techniques. Compared to 3DCRT, which has the lowest number of MU, 3.9 times higher. Hall et al. also found that IMRT has 2 to 3 times more MU compared to 3DCRT, which confirms our results. The mean value of the irradiation schedules with VMAT is about 370 MU, HYBT is exactly between 3DCRT and VMAT, as expected. The number of MU depends, among other things, on the size of the irradiation field. When examining the individual irradiation fields, the 3DCRT has the largest open fields; with the VMAT, despite MLC movement during irradiation, no long-lasting very small fields could be detected either. This could be seen from the observation of the “Beams-Eye-View-Simulations” and the “Control Points” of the MLC. Only with IMRT could small irradiation fields in the form of “slits” be detected due to the sliding window technique. This also explains the increased number of MU. Smaller fields and the associated higher MU numbers lead to an increased scattered radiation load, which originates from the gantry head. This leads to an increased low-dose exposure to a larger area of the body and thus also to a higher whole-body exposure, which promotes the risk of a second carcinoma. Hall and Wu describe radiotherapy as a double-edged sword; on the one hand it is used to treat cancer, but on the other hand, it can also cause cancer [14].

There is no influence of the **different breast groups** on the **number of MU** in 3DCRT, HYBT and VMAT. Only in IMRT,the G and Std groups have significantly more MU than K and Thxw. Despite the recommendation in the S3 guideline for breast carcinomas to use IMRT for large breast sizes, it appears that the number of MU is higher with IMRT for larger volumes such as G and Std. This may be related to the deeper or thicker irradiation volume, as longer irradiation is required to deliver more dose at depth.

The **homogeneity index** shows similar values on average for all four irradiation techniques. IMRT has the lowest HI and therefore the most homogeneous dose distribution. This irradiation technique is a pure fluence-based irradiation planning and with the existing tools in the irradiation planning, the most homogeneous dose possible is achieved in the target volume, which is also reflected in the results. Strongly modulated IMRT produces small irradiation fields with high MU. 3DCRT usually has large/open irradiation fields; depending on the number of subfields, a more homogeneous dose distribution can be achieved in the target volume. The generated 3DCRT treatment plans use few fields and result in relatively hard dose transitions, which lead to a more inhomogeneous dose distribution in the target volume. VMAT has more possibilities to distribute the dose in the irradiation volume due to the rotational irradiation and the moving MLC leaves. Depending on the modulation, VMAT also generates smallest fields, similar to IMRT. In this case, HYBT lies in the middle between 3DCRT and VMAT. As a result, the dose homogeneity depends on the number and modulation of the irradiation fields.

When looking at the individual breast groups, it is noticeable that breast group G has a more homogeneous distribution than the mean value of the individual irradiation techniques. For 3DCRT, this is probably due to the higher number of irradiation fields. With IMRT and VMAT, the treatment planning program has more modulation options due to the larger irradiation volume and can achieve a more homogeneous dose distribution. The group Thxw shows a better homogeneity as an exception with IMRT. The group Std has the most inhomogeneous dose distribution regardless of the irradiation technique, which is presumably also due to the number and modulation possibilities of the irradiation fields. Another approach would be the possible influence of the different breast shapes. Std are more conical/tubular compared to G. The groups G and K have a more square volume. The difference in the diameters of the tangential layers between the first mammillary layers and the posterior layers near the ribs is proportionally greater in the Std group than in the G and K groups; this means that there is a more inhomogeneous volume, which has an influence on the dose distribution.

### Conformity index

VMAT has the highest conformity because it has more possibilities for dose modulation. Techniques such as VMAT and IMRT, where the irradiation fields are only specified to a limited extent by the user, have a better CI. This can probably be attributed to the DVH optimization, as these techniques are inverse irradiation planning. 3DCRT has the lowest compliance, although it is on average only 0.07 worse than IMRT. The formula used considers the volume with the prescribed isodose outside the target volume, which explains the poor CI for 3DCRT. Due to the tangential irradiation fields used, part of the normal tissue next to the PTV is also irradiated in 3DCRT. Depending on the marked target volume, more or less normal tissue outside the PTV is also irradiated. The irradiated volume is usually fat and muscle, both of which are not considered high-risk organs. The breast volume is often not clearly distinguishable from normal tissue, so the question arises whether 3DCRT offers an additional safety margin by covering the areas outside the PTV.

The larger breast groups show a higher conformity for all irradiation techniques. With regard to the dose calculation of this planning program among the irradiation techniques used, it is noticeable that the dose under the skin is initially lower, which probably reflects the typical build-up effect of photon radiation. Smaller target volumes have a higher percentage of direct volume under the skin compared to larger volumes and this has a greater impact on conformity. Hence, the Thxw group in particular has a low conformation.

Considering the techniques used, it can be said that an appropriate radiation plan could be established with all of them. Nevertheless, there are several points that should be considered in more detail. The radiation planning is based on a computer tomography image, which represents a momentary image. Volumes in the thoracic region, in particular, are often in motion due to the patient’s natural breathing; this also applies to chest irradiation. In order to avoid partial areas not being irradiated or not being irradiated properly during breathing, different procedures were used in the techniques to minimize deficits caused by breathing. While in 3DCRT and IMRT a dose safety margin to the outside could be established by different techniques, no corresponding possibility could be found for VMAT. This could lead to undesired dose coverage during irradiation with thoracic movement. In addition, VMAT irradiates significantly more into the body than the other techniques, but predominantly in the low-dose range. In order to be able to make a more accurate statement, the irradiation plans created would have to be verified, for example, with a suitable thorax body phantom that simulates natural breathing. Dose measurements could then be taken, which would determine any over- or under-exposure of the irradiation volume. Due to the fact that VMAT is being used more and more frequently in breast cancer irradiation, the recurrence rates in breast carcinoma should be closely examined in the coming years depending on the irradiation technique.

The results of this study can be compared with previously conducted studies by Rastogie et al, Das Majumdar et al, Garg or Hall et al. Similar to the existing results regarding the radiation dose to organs at risk, it is also shown that 3DCRT proves to be beneficial in relation to low-dose areas. In contrast to these studies, additional attention is paid to the different breast groups. Due to the fact that similar results of the respective irradiation technique are achieved regardless of the breast group, it can be concluded that the size of the irradiated breast does not have a significant influence on the dose received.

Taking into account the results of this study, it can still be generally concluded that the quality of the treatment plan, regardless of the technique used, varies greatly depending on the experience of the physicist in question. Repeating the study with physicists from different institutes and with different age averages would most likely yield further interesting results. Furthermore, increasing the number of cases would lead to a statistically more reliable result. The irradiation techniques considered in this study were only compared on one system from one manufacturer. It would also be of interest to repeat this on other planning systems from other manufacturers, as these show clear differences. Another approach would be to examine whether an even lower-risk irradiation could be achieved by changing the patient positioning or using an alternative positioning system.

## 5. Conclusion

To evaluate the irradiation techniques for left-sided mammary irradiation, different parameters such as the OAR strains, the MU number, the HI and the CI were considered. Eighty treatment plans with different irradiation techniques were created and compared. Historically, 3DCRT has been the most widely used technique and it was also shown in this study that newer techniques are not necessarily better, even if they show a good result at first sight. With all the techniques used, adequate irradiation plans could be established. However, there are several issues that need to be considered when assessing the results. One of these is that the radiation planning is based on a computer tomography image and thus on a momentary image. In order to be able to make a better statement, the treatment plans created would have to be verified, for example, with a suitable thorax body phantom that simulates natural breathing. Additionally, since VMAT is being used more and more frequently in breast cancer irradiation, the recurrence rates of breast carcinoma depending on the irradiation technique used should be closely examined in the coming years.

We can partially confirm the recommendation of the S3 guideline for breast cancer that IMRT or VMAT should generally not be used for breast cancer radiotherapy in adjuvant radiotherapy, but only in the case of larger breasts and/or abnormal thoracic curvature [2]. In exceptional cases, IMRT can be helpful in the extreme sparing of individual OARs, with 3DCRT often being superior to IMRT in terms of mean cardiac dose [23]. The results are consistent with these findings; 3DCRT is still a very good irradiation technique for breast cancer irradiation. The study shows that a subdivision into different breast groups has no significant effect on the dose received.

In conclusion, due to the different level of experience of the physicist and the circumstances of the planning program used, there are certain influences on the results of this study. The study could therefore be repeated by other institutes and using different planning systems in order to provide a significant result and to verify the statements made in this study. In addition, a change in patient positioning should be considered to alter the shape of the breast and therefore better protect the OAR. A renewed evaluation of the various radiation techniques with regard to their suitability for the irradiation of breast carcinomas would then be necessary.

## Data Availability

All data produced in the present study are available upon reasonable request to the authors

## Litertaturverweise (References)

[1] Robert Koch-Institut und Gesellschaft Der Epidemiologischen Krebsregister In Deutschland E.V ., „Krebs in Deutschland 2015/2016”, Robert Koch-Institut, 2019. doi: 10.25646/5977.2.

[2] „Leitlinienprogramm Onkologie (Deutsche Krebsgesellschaft, Deutsche Krebshilfe, AWMF): S3-Leitlinie Früherkennung, Diagnose, Therapie und Nachsorge des Mammakarzinoms, Version 4.4, 2021, AWMF Registernummer: 032-045OL, http://www.leitlinienprogramm-onkologie.de/leitlinien/mammakarzinom/ (abgerufen am: 09.02.2024)”, 2021.

[3] M. Wannenmacher, F. Wenz, und J. Debus, Hrsg., Strahlentherapie. Berlin, Heidelberg: Springer Berlin Heidelberg, 2013. doi: 10.1007/978-3-540-88305-0.

[4] S. C. Lymberis u. a., „Prospective Assessment of Optimal Individual Position (Prone Versus Supine) for Breast Radiotherapy: Volumetric and Dosimetric Correlations in 100 Patients”, International Journal of Radiation Oncology*Biology*Physics, Bd. 84, Nr. 4, S. 902–909, Nov. 2012, doi: 10.1016/j.ijrobp.2012.01.040.

[5] Z. Varga, K. Hideghéty, T. Mező, A. Nikolényi, L. Thurzó, und Z. Kahán, „Individual Positioning: A Comparative Study of Adjuvant Breast Radiotherapy in the Prone Versus Supine Position”, International Journal of Radiation Oncology*Biology*Physics, Bd. 75, Nr. 1, S. 94–100, Sep. 2009, doi: 10.1016/j.ijrobp.2008.10.045.

[6] K. L. Griem, P. Fetherston, M. Kuznetsova, G. S. Foster, S. Shott, und J. Chu, „Three-dimensional photon dosimetry: a comparison of treatment of the intact breast in the supine and prone position”, International Journal of Radiation Oncology*Biology*Physics, Bd. 57, Nr. 3, S. 891–899, Nov. 2003, doi: 10.1016/S0360-3016(03)00723-5.

[7] E.-O. O. Osa u. a., „Prone Breast Intensity Modulated Radiation Therapy: 5-Year Results”, International Journal of Radiation Oncology*Biology*Physics, Bd. 89, Nr. 4, S. 899–906, Juli 2014, doi: 10.1016/j.ijrobp.2014.03.036.

[8] S. C. Darby, P. McGale, C. W. Taylor, und R. Peto, „Long-term mortality from heart disease and lung cancer after radiotherapy for early breast cancer: prospective cohort study of about 300 000 women in US SEER cancer registries”, The Lancet Oncology, Bd. 6, Nr. 8, S. 557–565, Aug. 2005, doi: 10.1016/S1470-2045(05)70251-5.

[9] M. D. Piroth u. a., „Heart toxicity from breast cancer radiotherapy: Current findings, assessment, and prevention”, Strahlenther Onkol, Bd. 195, Nr. 1, S. 1–12, Jan. 2019, doi: 10.1007/s00066-018-1378-z.

[10] M. Stovall u. a., „Dose to the Contralateral Breast From Radiotherapy and Risk of Second Primary Breast Cancer in the WECARE Study”, International Journal of Radiation Oncology*Biology*Physics, Bd. 72, Nr. 4, Art. Nr. 4, Nov. 2008, doi: 10.1016/j.ijrobp.2008.02.040.

[11] K. Rastogi, S. Sharma, S. Gupta, N. Agarwal, S. Bhaskar, und S. Jain, „Dosimetric comparison of IMRT versus 3DCRT for post-mastectomy chest wall irradiation”, Radiat Oncol J, Bd. 36, Nr. 1, S. 71–78, März 2018, doi: 10.3857/roj.2017.00381.

[12] S. K. Das Majumdar u. a., „A Dosimetric Study Comparing 3D-CRT vs. IMRT vs. VMAT in Left-Sided Breast Cancer Patients After Mastectomy at a Tertiary Care Centre in Eastern India”, Cureus, März 2022, doi: 10.7759/cureus.23568.

[13] A. Garg und P. Kumar, „Dosimetric Comparison of the Heart and Left Anterior Descending Artery in Patients With Left Breast Cancer Treated With Three-Dimensional Conformal and Intensity-Modulated Radiotherapy”, Cureus, Jan. 2022, doi: 10.7759/cureus.21108.

[14] E.J. Hallund C.-S. Wuu, „Radiation-induced second cancers: the impact of 3D-CRT and IMRT”, International Journal of Radiation Oncology*Biology*Physics, Bd. 56, Nr. 1, Art. Nr. 1, Mai 2003, doi: 10.1016/S0360-3016(03)00073-7.

[15] M. B. Mukesh u. a., „Randomized Controlled Trial of Intensity-Modulated Radiotherapy for Early Breast Cancer: 5-Year Results Confirm Superior Overall Cosmesis”, JCO, Bd. 31, Nr. 36, S. 4488–4495, Dez. 2013, doi: 10.1200/JCO.2013.49.7842.

[16] B. McCormick und M. Hunt, „Intensity-Modulated Radiation Therapy for Breast: Is It for Everyone?”, Seminars in Radiation Oncology, Bd. 21, Nr. 1, S. 51–54, Jan. 2011, doi: 10.1016/j.semradonc.2010.08.009.

[17] F. Lohr u. a., „Ist die Kardiotoxizität der Radiotherapie im Rahmen des Brusterhalts überhaupt noch relevant, und könnte sie durch Mehrfelder-IMRT gesenkt werden?”, Strahlenther Onkol, Bd. 185, Nr. 4, S. 222–230, Apr. 2009, doi: 10.1007/s00066-009-1892-0.

[18] F. R. Bartlett u. a., „The UK HeartSpare Study (Stage IB): Randomised comparison of a voluntary breath-hold technique and prone radiotherapy after breast conserving surgery”, Radiotherapy and Oncology, Bd. 114, Nr. 1, S. 66–72, Jan. 2015, doi: 10.1016/j.radonc.2014.11.018.

[19] S. Stroh, M. Even, R. Berghs, J. Trzewik, und C. Stegmann, „HYBT-A combined radiation technique for breast cancer using the EclipseTM version 15.6 planning system from Varian Medical Systems. YRA–Young Researchers Academy MedTech in NRW, 13”, 2024.

[20] M. Aghili, M. Barzegartahamtan, A. Alikhassi, und R. Mohammadpour, „Investigation of electron boost radiotherapy in patients with breast cancer: Is a direct electron field optimal?”, Cancer/Radiothérapie, Bd. 22, Nr. 1, S. 52–56, Feb. 2018, doi: 10.1016/j.canrad.2017.08.109.

[21] H. Van Parijs, T. Reynders, K. Heuninckx, D. Verellen, G. Storme, und M. De Ridder, „Breast conserving treatment for breast cancer: dosimetric comparison of different non-invasive techniques for additional boost delivery”, Radiat Oncol, Bd. 9, Nr. 1, S. 36, Dez. 2014, doi: 10.1186/1748-717X-9-36.

[22] J. Unnithan und R.M. Macklis, „Contralateral breast cancer risk”, Radiotherapy and Oncology, Bd. 60, Nr. 3, S. 239–246, Sep. 2001, doi: 10.1016/S0167-8140(01)00369-3.

[23] „Abstracts DEGRO 2016”, Strahlenther Onkol, Bd. 192, Nr. S1, Art. Nr. S1, Juni 2016, doi: 10.1007/s00066-016-0974-z.

